# Genetic and potential antigenic evolution of influenza A(H1N1)pdm09 viruses circulating in Kenya during 2009-2018 influenza seasons

**DOI:** 10.1101/2022.04.13.22273796

**Authors:** D. Collins Owuor, Zaydah R. de Laurent, Bryan O. Nyawanda, Gideon O. Emukule, Rebecca Kondor, John R. Barnes, D. James Nokes, Charles N. Agoti, Sandra S. Chaves

## Abstract

**Background:** Influenza viruses undergo rapid evolutionary changes, which requires continuous surveillance to monitor for genetic and potential antigenic changes in circulating viruses that can guide control and prevention decision making.

**Methods:** We sequenced and phylogenetically analyzed A(H1N1)pdm09 virus genome sequences obtained from specimens collected from hospitalized patients of all ages with or without pneumonia between 2009 and 2018 from seven sentinel surveillance sites across Kenya. We compared these sequences with recommended vaccine strains during the study period to infer genetic and potential antigenic changes in circulating viruses and determinants of clinical outcome.

**Results:** We generated and analyzed a total of 383 A(H1N1)pdm09 virus genome sequences. Phylogenetic analyses revealed that multiple genetic groups (clades, subclades, and subgroups) of A(H1N1)pdm09 virus circulated in Kenya over the study period; these evolved away from their vaccine strain, forming clades 7 and 6, subclades 6C, 6B, and 6B.1, and subgroups 6B.1A and 6B.1A1. Several amino acid substitutions among circulating viruses were associated with continued evolution of the viruses, especially in antigenic epitopes and receptor binding sites (RBS) of circulating viruses. Disease severity reduced with increase in age among children aged <5 years.

**Conclusion:** Our study highlights the utility of genomic surveillance to monitor the evolutionary changes of influenza viruses. Routine influenza surveillance with broad geographic representation and whole genome sequencing capacity to inform on the severity of circulating strains could improve selection of influenza strains for inclusion in vaccines.

## Introduction

The first pandemic of the 21^st^ century was a result of an influenza A virus (IAV) designated A(H1N1)pdm09 virus, which emerged in North America during March-April 2009 and spread rapidly among humans [1-3]. A(H1N1)pdm09 virus displaced seasonal A(H1N1) virus and has continued to circulate in subsequent years alongside influenza A(H3N2) and B viruses globally [4-6], causing annual seasonal epidemics [7-9]. The rapid ability of the virus to spread globally upon its emergence highlights the public health threat posed by emergent viruses [3, 10, 11]. The countrywide surveillance for influenza viruses among medically-attended patients in Kenya since the emergence of the A(H1N1)pdm09 virus strain allows for exploring the phenotypic characteristics of the virus [7, 12, 13].

We characterized the genetic and antigenic evolution in A(H1N1)pdm09 viruses circulating in Kenya between 2009 and 2018 using codon-complete gene sequences generated through next-generation sequencing (NGS). We then utilized genetic sequence data and available clinical information to investigate determinants of clinical outcome among hospitalized children aged <5 years.

## Materials and Methods

### Study design

Samples analyzed in this study were collected between June 2009 and December 2018 through two health facility-based surveillance networks in Kenya as detailed previously [13]. The first involved continuous countrywide surveillance for influenza through severe acute respiratory illness (SARI) sentinel hospital reporting undertaken at six sites supported by the Centers for Disease Control (CDC)-Kenya Country office: Kenyatta National Hospital (KNH), Nakuru County and Referral Hospital (CRH), Nyeri CRH, Kakamega CRH, Siaya CRH, and Coast General Teaching and Referral Hospital [7, 8, 13-16]. The second was the pediatric viral pneumonia surveillance undertaken at Kilifi County Hospital (KCH) [12].

In the CDC-supported surveillance sites (CDC-Kenya), SARI was defined as acute onset of illness (within the last 14 days) among hospitalized patients of all ages with cough and reported fever (feverish) or a recorded temperature of ≥38°C. Demographics, underlying diseases, and signs and symptoms were collected from patients who met the case definition; the patients were also assessed by study clinicians on physical and clinical findings. Furthermore, among hospitalized children aged <5 years, additional clinical information including difficulty in breathing, lower chest wall indrawing, inability to drink or breastfeed, and nasal flaring were recorded. Chart review was done at the time of discharge or death to collect clinical outcome data. In the second facility-based surveillance undertaken at KCH from January 2009 through December 2018, a definition of pediatric viral pneumonia among children aged 1 day to 59 months presenting with syndromic severe or very severe pneumonia was used. A history of cough for <30 days or difficulty breathing, when accompanied by lower chest wall indrawing was defined as severe pneumonia; a history of cough for <30 days or difficulty breathing, when accompanied by any one of prostration (including inability to feed or drink), coma, or hypoxemia (oxygen saturation <90%) was defined as very severe pneumonia [7, 16].

We selected a total of 418 (31.9%) A(H1N1)pdm09 virus positive SARI samples for this analysis based on real-time reverse-transcription (RT)-PCR cycle threshold (Ct) of <35.0 (as proxy for high viral load), adequate sample volume for RNA extraction (>140 μL), and balanced distribution of samples based on surveillance sites and years. We also identified a total of 157 IAV positive specimens from the KCH surveillance site. These were not subsequently subtyped for influenza A(H1N1)pdm09 and A(H3N2) viruses. Therefore, we utilized all these specimens in the current analysis.

### RNA extraction and multi-segment real-time PCR (M-RTPCR) for IAV

We performed viral nucleic acid extraction from IAV and A(H1N1)pdm09 virus positive samples using the QIAamp Viral RNA Mini Kit (Qiagen). We then reverse transcribed the extracted RNA, and amplified the complete coding region of IAV genome in a single M-RTPCR using the Uni/Inf primer set [17]. We evaluated successful amplification by running the products on 2% agarose gel and visualized the reaction on a UV transilluminator after staining with RedSafe Nucleic Acid Staining solution (iNtRON Biotechnology Inc.,).

### IAV NGS and virus genome assembly

Following PCR, we purified the amplicons with 1X AMPure XP beads (Beckman Coulter Inc., Brea, CA, USA), quantified amplicons with Quant-iT dsDNA High Sensitivity Assay (Invitrogen, Carlsbard, CA, USA), and normalized amplicons to 0.2 ng/μL. We generated indexed paired-end libraries from 2.5 μL of 0.2 ng/μL amplicon pool using Nextera XT Sample Preparation Kit (Illumina, San Diego, CA, USA) following the manufacturer’s protocol. We then purified amplified libraries using 0.8X AMPure XP beads, quantitated libraries using Quant-iT dsDNA High Sensitivity Assay (Invitrogen, Carlsbard, CA, USA), and evaluated libraries for fragment size in the Agilent 2100 BioAnalyzer System using the Agilent High Sensitivity DNA Kit (Agilent Technologies, Santa Clara, CA, USA). We diluted the libraries to 2nM in preparation for pooling and denaturation for running on the Illumina MiSeq (Illumina, San Diego, CA, USA). We then NaOH denatured pooled libraries, diluted libraries to 12.5 pM, and sequenced on the Illumina MiSeq using 2 × 250 bp paired end reads with the MiSeq v2 500 cycle kit (Illumina, San Diego, CA, USA). We added five percent Phi-X (Illumina, San Diego, CA, USA) spike-in to the libraries to increase library diversity by creating a more diverse set of library clusters. We carried out contiguous (contigs) nucleotide sequence assembly from the sequence data using the FLU module of the Iterative Refinement Meta-Assembler (IRMA) using IRMA default settings. We deposited all the generated sequence data in the Global Initiative on Sharing All Influenza Data (GISAID) EpiFlu™ database (https://platform.gisaid.org/epi3/cfrontend).

### Phylogenetic clustering and genetic group classification

We aligned and translated consensus nucleotide sequences for all gene segments in AliView version 1.26 (https://ormbunkar.se/aliview/). We then reconstructed Maximum-likelihood (ML) trees for the individual gene segments using IQ-TREE version 2.0.7 (http://www.iqtree.org/). The software initiates tree reconstruction after assessment and selection of the best model of nucleotide substitution for alignment. We linked the ML trees to various metadata and visualized using R ggtree version 2.4.2 in R programming software v4.0.2 (http://www.rstudio.com/). We used the codon-complete hemagglutinin (HA) sequences of all viruses to characterize A(H1N1)pdm09 virus strains into genetic groups (i.e., clades, subclades, and subgroups) using Phylogenetic Clustering using Linear Integer Programming (PhyCLIP) v2.0 (https://github.com/alvinxhan/PhyCLIP). We downloaded vaccine strains and reference viruses from GISAID EpiFlu™ database (https://platform.gisaid.org/epi3/cfrontend). We aligned and reconstructed ML trees using vaccine strains, reference viruses, and translated Kenyan A(H1N1)pdm09 viruses.

### Genetic characterization of amino acid substitutions in surface glycoproteins, matrix, and non-structural proteins

We aligned the hemagglutinin (HA) and neuraminidase (NA) glycoproteins, matrix (M2), and non-structural 1 (NS1) gene segments of A(H1N1)pdm09 virus from this study using MAFFT version 7.475 (https://mafft.cbrc.jp/alignment/software/). We identified the amino acid substitutions in the antigenic epitopes (Sa, Sb, Ca1, Ca2 and Cb) [18], receptor binding sites (RBS), and potential glycosylation sites of HA1 subunit of HA protein and substitutions in the codon-complete HA, NA, M2, and NS1 protein sequences of A(H1N1)pdm09 viruses using Flusurver (https://flusurver.bii.a-star.edu.sg; accessed on June 01, 2021). We generated highlighter plots of Kenyan and reference sequences (obtained from the Global Initiative on Sharing All Influenza Data (GISAID) EpiFlu™ database - https://platform.gisaid.org/epi3/cfrontend) using inhouse python scripts to visualize amino acid differences in M2 and NS1 protein sequences among the sampled viruses.

### Predictors of severe infection among hospitalized children aged <5 years

We used multivariable logistic regression in Stata version 16 (Stata Corp, College Station, Texas, USA) to investigate the predictors of severe infection among hospitalized patients. Only samples collected from hospitalized children aged <5 years from CDC-Kenya and KCH surveillance were used to estimate the predictors of severe infection. Children hospitalized with fever and acute cough were categorized as severe if they had breathing difficulty and/or lower chest wall indrawing, or otherwise non-severe. The predictors investigated included patient age (categorized as <12 months, 12-23 months, and ≥24 months), location of surveillance sites, year of A(H1N1)pdm09 virus sampling (pandemic period, 2009-10; post pandemic period, 2011 onwards), A(H1N1)pdm09 virus genetic group, antigenic epitope substitutions, NS1 protein substitutions, and Ct values as proxy for viral load distributed in tertiles.

### Ethics

Ethical clearance for the study was granted by the Kenya Medical Research Institute (KEMRI) Scientific Steering Committee (SSC# 1899, 2558, and 2692) and KEMRI - Wellcome Trust Research Programme Scientific and Ethical Review Unit (SERU# 1055, 1433, and 3044). Informed consent was sought and received from the study participants for the study.

## Results

### IAV sequencing and genome assembly

We generated and analyzed a total of 383 A(H1N1)pdm09 virus genome sequences. Among 418 A(H1N1)pdm09 virus positive samples from the CDC-Kenya surveillance system, 414 (99.1%) passed pre-sequencing quality control checks, which generated 344 (83.1%) codon-complete A(H1N1)pdm09 virus genome sequences on the MiSeq. Of the 157 IAV positive specimens available from KCH, 94 (59.9%) passed pre-sequencing quality control checks generating 45 (47.9%) A(H1N1)pdm09 virus (39 codon-complete and 6 partial) and 49 (52.1%) A(H3N2) virus genome sequences (46 codon-complete and 3 partial). For this report, only the 39 codon-complete A(H1N1)pdm09 virus sequences were included in the analyses. The sociodemographic and clinical characteristics of these patients are shown in **Table 1**.

**Table 1.** Sociodemographic and clinical characteristics of hospitalized patients in severe acute respiratory illness (SARI) and viral pneumonia surveillances in Kenya, 2009-18.

|  |  | SARI surveillance |  | Viral pneumonia surveillance |  |
| --- | --- | --- | --- | --- | --- |
|  |  | n | % | n | % |
| Age | 0-11 months | 73 | 21.2 | 18 | 46.2 |
|  | 12-59 months | 203 | 59.0 | 21 | 53.8 |
|  | >=5 years | 68 | 19.8 |  |  |
| Gender | Female | 141 | 41.0 | 14 | 35.9 |
|  | Male | 203 | 59.0 | 25 | 64.1 |
| Fever | No |  |  | 1 | 2.6 |
|  | Yes | 344 | 100.0 | 38 | 97.4 |
| Cough | No |  |  | 1 | 2.6 |
|  | Yes | 344 | 100.0 | 38 | 97.4 |
| Breathing difficulty | No | 244 | 70.9 | 2 | 5.1 |
|  | Yes | 100 | 29.1 | 37 | 94.9 |
| In-drawing‡ | No | 144 | 41.9 | 1 | 2.6 |
|  | Yes | 83 | 24.1 | 38 | 97.4 |
| Nasal flaring‡ | No | 158 | 45.9 | 13 | 33.3 |
|  | Yes | 82 | 23.8 | 26 | 66.7 |
| Unable to drink/breastfeed‡ | No | 188 | 54.7 | 4 | 10.3 |
|  | Yes | 26 | 7.6 | 35 | 89.7 |
| Lethargic‡ | No | 141 | 41.0 | 4 | 10.3 |
|  | Yes | 89 | 25.9 | 35 | 89.7 |
| Wheezing | No | 133 | 38.7 | 4 | 10.3 |
|  | Yes | 10 | 2.9 | 35 | 89.7 |
| Oxygen saturation | <90 | 29 | 8.4 | 6 | 15.4 |
|  | >=90 | 128 | 37.2 | 33 | 84.6 |
| Intensive Care Unit | No | 63 | 18.3 |  |  |
|  | Yes | 15 | 4.4 |  |  |
| Outcome | Death | 8 | 2.3 |  |  |
|  | Discharged | 176 | 51.2 |  |  |
|  | Absconded | 4 | 1.2 |  |  |

‡ Chest in-drawing, nasal flaring, unable to drink/breastfeed at all, and lethargy in patients aged <5 years in SARI surveillance. SARI, severe acute respiratory illness.

### Circulation of A(H1N1)pdm09 virus strains in Kenya and their patterns of antigenic drift

Phylogenetic analyses revealed that multiple genetic groups of influenza A(H1N1)pdm09 virus circulated in Kenya over the study period and evolved away from their vaccine strain, **Figure 1**. All Kenyan viruses from 2009-18 evolved away from the vaccine strains A/California/07/2009 (H1N1pdm09)-like virus and A/Michigan/45/2015 (H1N1pdm09)-like virus. The A(H1N1)pdm09 virus strains from 2009 to 2016 (n=324; 84.6%) evolved away from the 2009-17 Northern Hemisphere (NH) and 2009-16 Southern Hemisphere (SH) vaccine strain A/California/07/2009 (H1N1pdm09)-like virus and fell into clade 7 (n=97, 25.3%), clade 6 (n=132, 34.5%), subclade 6C (n=10, 2.6%), subclade 6B (n=47, 12.3%), and subclade 6B.1 (n=38, 9.9%), respectively. All the viruses from 2018 evolved away from the 2017-18 NH and 2017 SH vaccine strain A/Michigan/45/2015 (H1N1pdm09)-like virus and fell into subgroup 6B.1A (n=57, 14.9%) and subgroup 6B.1A1 (n=2, 0.5%), respectively.

**Figure 1.**
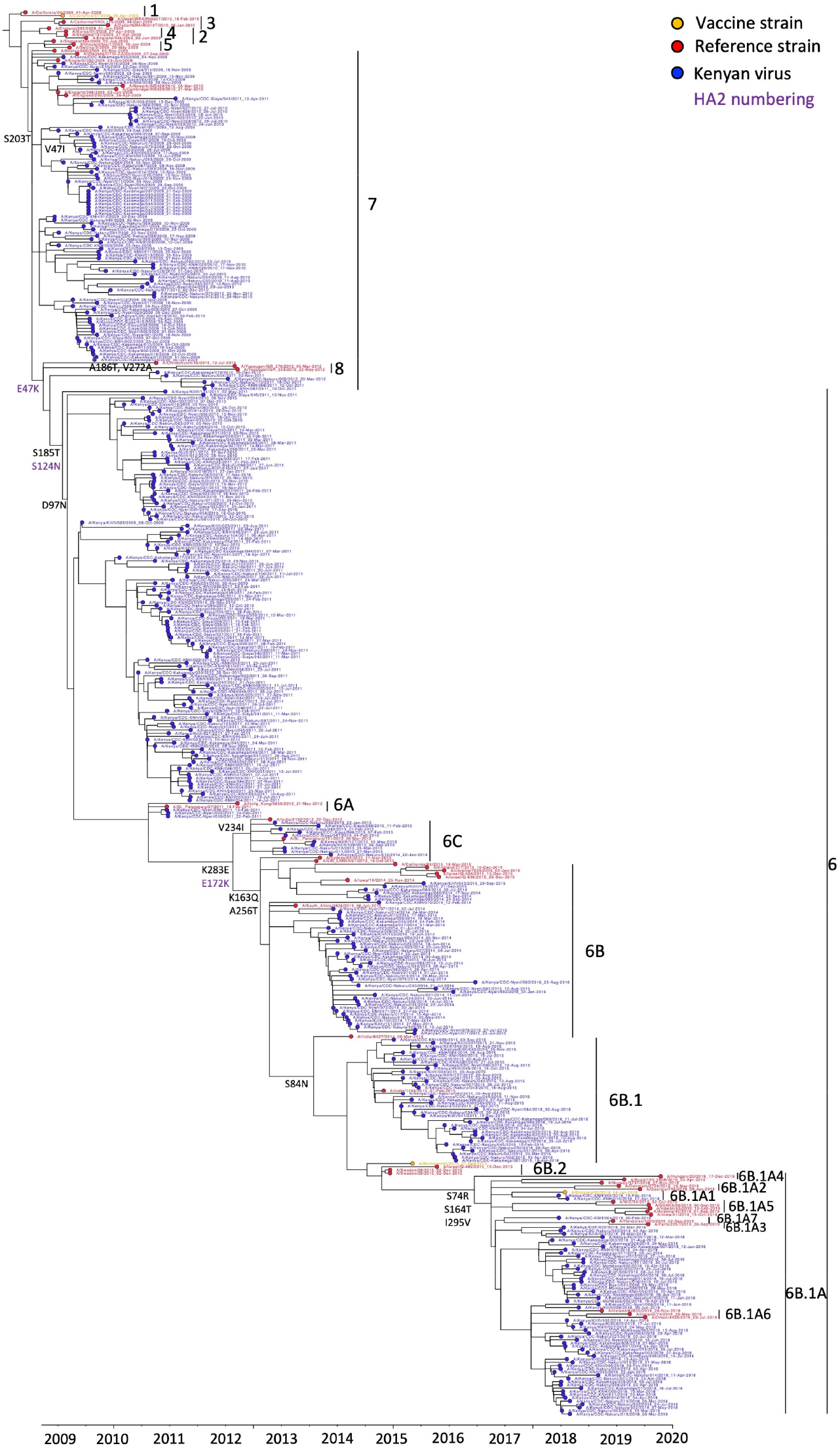
Maximum-likelihood phylogenetic tree of A(H1N1)pdm09 virus sequences from Kenya collected between 2009 and 2018, A(H1N1)pdm09 virus vaccine strains, and reference viruses. This is a time-calibrated phylogenetic tree with time shown on the x-axis. Branches are colored based on Kenyan, vaccine, and reference strains as shown in the color key. Amino acid substitutions in HA1 and HA2 subunits, which define A(H1N1)pdm09 virus genetic groups are also shown (HA2 substitutions are labelled in purple as shown in the color key).

Clades 6 and 7 viruses co-circulated in Kenya between 2009 and 2012, with clade 6 viruses carrying HA substitutions D97N, S185T, and S203T, while clade 7 viruses carried HA substitutions D97, S185, and S203T, **Figure 1**. Clade 6 viruses evolved into subclades 6A, 6B, and 6C; 6C carrying HA substitution V234I circulated in Kenya in 2013-14, while 6B with HA substitutions K163Q, A256T, and K283E circulated in Kenya in 2014-16. Subclade 6B evolved further into subgroup 6B.1 carrying additional HA substitution S84N, which dominated in 2015-16. Subgroup 6B.1 further evolved into subgroups 6B.1A and 6B.1A1 carrying HA substitutions S74R, S164T, and I295V that dominated in 2018.

Comparison of the deduced amino acid sequences of the viruses identified in hospitalized patients relative to the vaccine strains from which all the 2009-18 strains considerably evolved revealed significant amino acid substitutions in antigenic epitopes and RBS among the circulating A(H1N1)pdm09 virus strains (**Table 2**). There were 13 amino acid substitutions across the five antigenic sites among sampled Kenyan viruses. Ranking from the most variable site, Sa, Ca2, Sb, Ca1, and Cb had two, six, six, three, and one substitution, respectively. The most frequent HA substitution per site for Sa, Sb, Ca1, Ca2 and Cb was K163Q, S185T, S203T, A141E, and S74R, respectively. HA substitutions S203T and S185T were the most dominant substitutions occurring in 100% and 59.3% (227/383) viruses from 2009-18, respectively. In addition, we observed previously described HA substitutions at the RBS (A186T and D222E) and glycosylation sites (A186T and N125S) of the viruses.

**Table 2.** Antigenic drift among A(H1N1)pdm09 virus strains collected from Kenya, 2009-18.

| Season | Vaccine (Genetic group); hemisphere ‡ | Number of Kenyan viruses analyzed (year of sampling) | Amino acid substitutions † |
| --- | --- | --- | --- |
| 2009-2017 | A/California/07/2009 (clade 1); NH | 324 (2009-2016) | P137S<sup>d</sup>, A139V<sup>d</sup>, A141T<sup>d</sup>, K142R<sup>d</sup>, K163Q<sup>a</sup>, G170R<sup>c</sup>, S185T<sup>b</sup>, S185A<sup>b</sup>, S185I<sup>b</sup>, A186T<sup>b</sup>, A186V<sup>b</sup>, D187N<sup>b</sup>, S203T<sup>c</sup>, R205K<sup>c</sup>, D222E, D222N |
| 2009-2016 | A/California/07/2009 (clade 1); SH | 324 (2009-2016) | P137S<sup>d</sup>, A139V<sup>d</sup>, A141T<sup>d</sup>, K142R<sup>d</sup>, K163Q<sup>a</sup>, G170R<sup>c</sup>, S185T<sup>b</sup>, S185A<sup>b</sup>, S185I<sup>b</sup>, A186T<sup>b</sup>, A186V<sup>b</sup>, D187N<sup>b</sup>, S203T<sup>c</sup>, R205K<sup>c</sup>, D222E, D222N |
| 2017-2018 | A/Michigan/45/2015 (6B.1); NH | 59 (2018) | S74R<sup>e</sup>, P137S<sup>d</sup>, A141E<sup>d</sup>, S164T<sup>a</sup> |
| 2017 | A/Michigan/45/2015 (6B.1); SH | 59 (2018) | S74R<sup>e</sup>, P137S<sup>d</sup>, A141E<sup>d</sup>, S164T<sup>a</sup> |

NH, Northern Hemisphere; SH, Southern Hemisphere.
† A (H1N1)pdm09 antigenic sites are represented as Sa-a; Sb-b; Ca<sub>1</sub>-c; Ca<sub>2</sub>-d; and Cb-e.
‡ Recommended vaccines for each influenza season are adopted from Global Initiative on Sharing All Influenza Data (GISAIID) (https://www.gisaid.org/resources/human-influenza-vaccine-composition/).

### Genetic and antigenic characterization of NA, M2, and NS1 proteins

All NA proteins lacked the H275Y marker associated with resistance to neuraminidase inhibitors. However, seven (1.8%) NA proteins had substitutions T362I (3), I117M (3), and V234I (1). Additionally, all the Kenyan viruses contained the adamantine-resistance marker S31N, **Figure 2**. All 59 viruses from 2018 had six amino acid substitutions (E55K, L90I, I123V, E125D, K131E, and N205S) in NS1 protein, **Figure 3**.

**Figure 2.**
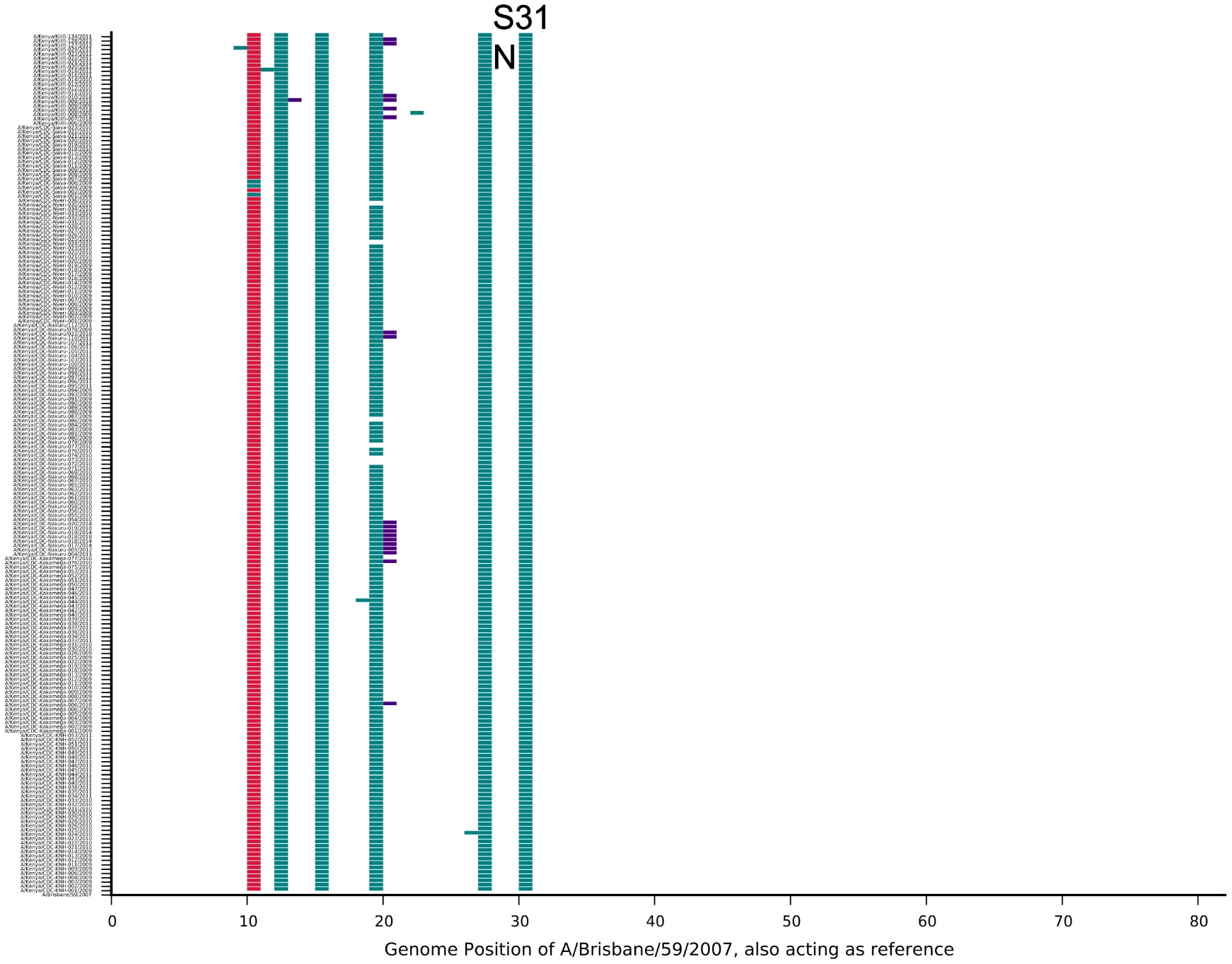
Highlighter plot of M2 protein sequences of A(H1N1)pdm09 viruses from Kenya showing the adamantine-resistance marker S31N amino acid substitution in M2 protein. The substitution was observed in all the A(H1N1)pdm09 viruses collected in Kenya between 2009 and 2018. Sequences were aligned with A/Brisbane/59/2007.

**Figure 3.**
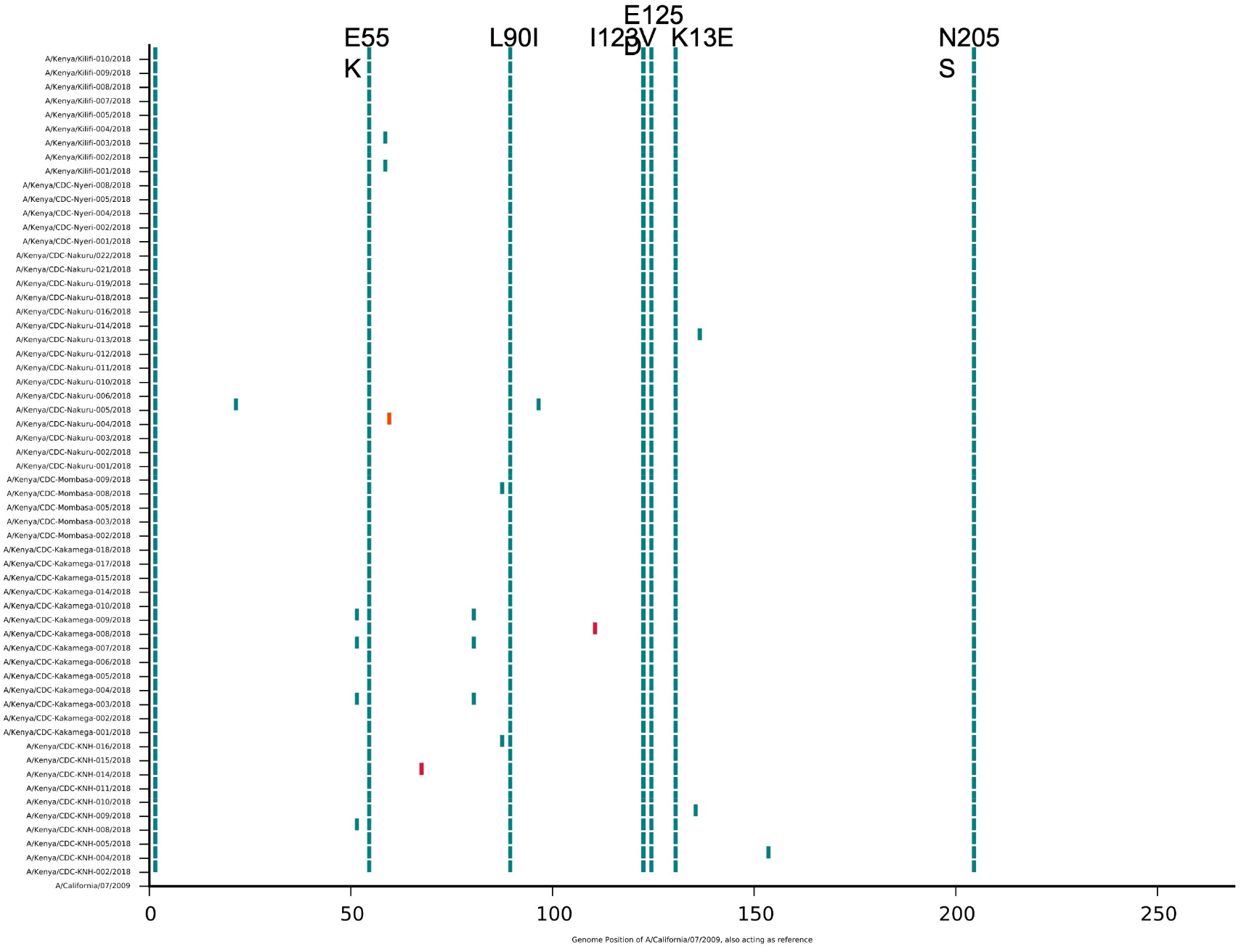
Highlighter plot of NS1 protein sequences of A(H1N1)pdm09 viruses from Kenya showing the NS1 protein substitutions: E55K, L90I, I123V, E125D, K131E, and N205S. The sequences were aligned with vaccine strain A/California/07/2009. The substitutions were reported for 59 virus sequences collected from hospitalized patients in 2018 from CDC-supported surveillance and KCH pediatric viral pneumonia surveillance. CDC, Centers for Disease Control; KCH, Kilifi County Hospital.

### Predictors of severe infection among hospitalized children aged <5 years

We observed that severity of infection reduced with increase in age. Children aged ≥24 months were less likely to have severe infection compared to children aged 0-11 months (adjusted odds ratio (aOR), 0.25; 95% CI, 0.11-0.54) and that A(H1N1)pdm09 viruses of subclade 6B and clade 7 were less likely associated with severe infection compared to clade 6 viruses (aOR, 0.25; 95% CI, 0.09-0.72) and (aOR, 0.34; 95% CI, 0.15-0.79), respectively (**Table 3**). Ct value, year of sampling, location of sampling, antigenic epitope substitutions, and NS1 protein substitutions were not associated with severe infection.

**Table 3.** Predictors of severe infection among hospitalized children aged <5 years.

| Characteristics | Severe, N=161<br>n (%) | Non-severe, N=108<br>n (%) | aOR (95% CI) | p-value |
| --- | --- | --- | --- | --- |
| Age |  |  |  |  |
| <12 months | 68 (42.2) | 20 (18.5) | Ref |  |
| 12-23 months | 58 (36.0) | 29 (25.9) | 0.61<br>(0.28,1.32) | 0.21 |
| ≥24 months | 35 (21.7) | 59 (54.6) | 0.25<br>(0.11,0.54) | <0.01 |
| Ct value (tertiles) |  |  |  |  |
| <22.5 | 42 (26.1) | 42 (38.9) | Ref |  |
| 22.5-25.5 | 46 (28.6) | 41 (38.0) | 0.87 (0.40,1.87) |  |
| >25.5 | 73 (45.3) | 25 (23.2) | 1.64 (0.76,3.53) |  |
| Year |  |  |  |  |
| Pandemic | 44 (27.3) | 32 (29.6) |  |  |
| Post-pandemic | 117 (72.7) | 76 (70.4) |  |  |
| Location |  |  |  |  |
| Kakamega | 15 (9.3) | 21 (19.4) | Ref |  |
| Kilifi | 39 (24.2) | 0 (0) | - |  |
| Mombasa | 5 (3.1) | 0 (0) | - |  |
| Nairobi | 33 (20.5) | 19 (17.6) | 1.60<br>(0.56,4.60) | 0.38 |
| Nakuru | 36 (22.4) | 38 (35.2) | 1.38<br>(0.54,3.56) | 0.5 |
| Nyeri | 18 (11.2) | 22 (20.4) | 1.78<br>(0.61,5.22) | 0.29 |
| Siaya | 15 (9.3) | 8 (7.4) | 2.01<br>(0.55,7.31) | 0.29 |
| Genetic group |  |  |  |  |
| Clade 6 | 55 (34.2) | 23 (21.3) | Ref |  |
| Subclade 6B | 14 (8.7) | 25 (23.2) | 0.25<br>(0.09,0.72) | 0.01 |
| Subclade 6B.1 | 32 (19.9) | 5 (4.63) | 2.68<br>(0.81,8.94) | 0.11 |
| Subgroup 6B.1A | 31 (19.3) | 21 (19.4) | 0.62<br>(0.24,1.58) | 0.31 |
| Subgroup 6B.1A1 | 2 (1.2) | 0 (0) |  |  |
| Subclade 6C | 4 (2.5) | 2 (1.9) | 0.68<br>(0.08,5.90) | 0.72 |
| Clade 7 | 23 (14.3) | 32 (29.6) | 0.34<br>(0.15,0.79) | 0.01 |
| Antigenic substitution |  |  |  |  |
| Yes | 99 (61.5) | 70 (64.8) |  |  |
| No | 62 (38.5) | 38 (35.2) |  |  |
| NS1 gene substitution |  |  |  |  |
| Yes | 33 (20.5) | 21 (19.4) |  |  |
| No | 128 (79.5) | 87 (80.6) |  |  |

aOR, adjusted odds ratio; CI, confidence interval; Ct; cycle threshold.

## Discussion

We observed that A(H1N1)pdm09 viruses circulating in Kenya from 2009-18 evolved away from their vaccine strain over the study period. Kenyan virus strains from 2009-16 evolved away from the 2009-17 NH and 2009-16 SH vaccine strain A/California/07/2009 (H1N1pdm09)-like virus and fell into clades 7 and 6, and subclades 6C, 6B, and 6B.1, respectively. All viruses from 2018 evolved away from the 2017-18 NH and 2017 SH vaccine strain A/Michigan/45/2015 (H1N1pdm09)-like virus and fell into subgroups 6B.1A and 6B.1A1, respectively. We identified considerable amino acid substitutions in antigenic epitopes and RBS among the circulating viruses, which confirms the continued evolution of circulating influenza viruses in Kenya. We recently reported that the evolutionary dynamics of A(H1N1)pdm09 virus in Kenya was associated with multiple virus introductions into the country between 2009 and 2018, although only a few of those introductions instigated local seasonal epidemics that then established local transmission clusters across the country [13]. We also observed substitutions in NA protein, which have been associated with reduced susceptibility to neuraminidase inhibitors in vitro [19], substitutions in M2 protein associated with adamantine-resistance, and substitutions in NS1 protein that possibly result in increased virulence [20]. Nonetheless, we were not able to associate viral genetic changes and substitutions with increased severity.

Analysis of virus sequence data from Kenya during the pandemic in 2009 identified the introduction of clades 2 and 7 viruses into Kenya [21]. We recently confirmed these findings and showed that clades 6 and 7 viruses were introduced into Kenya, disseminated countrywide, and persisted across multiple epidemics as local transmission clusters [13]. A recent study from Kenya reported the circulation of clade 6B, subclade 6B.1, and subclade 6B.2 viruses in the 2015-18 influenza seasons [22]. Here, through detailed genomic analysis, we extend these observations and show that multiple influenza strains were introduced into Kenya and spread countrywide over the study period. Most of the amino acid substitutions associated with the continued evolution of A(H1N1)pdm09 viruses in Kenya have also been reported in other studies in Africa [22-25] and Asia [26, 27]. Therefore, the continuing evolution of A(H1N1)pdm09 viruses in Kenya is in part due to the global circulation of influenza viruses.

The genetic diversity of A(H1N1)pdm09 viruses in specific regions arising from multiple virus introductions and subsequent establishment of local transmission clusters [13] composed of viruses harboring considerable amino acid substitutions in antigenic epitopes and RBS could lead to predominance of circulating viruses that are not closely matched to previously selected vaccine strains as shown in our study. In countries like Kenya, where influenza virus spread is year-round [9], there also exists unpredictability of which genetic virus strain may predominate and when. Influenza vaccines with broad coverage (“universal” vaccines) could be key to managing the influenza disease burden in such settings. Currently, Kenya does not have a national influenza vaccination policy [28], but it would be important to consider deployment of quadrivalent influenza vaccines with representative A(H1N1)pdm09 virus, A(H3N2) virus, and both influenza B virus lineages (B/Victoria and B/Yamagata) for optimal vaccine effectiveness [13, 29, 30]. It will also be important to investigate further whether the use of SH or NH formulated vaccines could have a place in tropical regions like in Kenya, where virus importations from both hemispheres are common.

Specific influenza virus gene segment phylogenies and genetic group memberships have been associated with disease severity [31]. Although we did not observe associations between genetic group membership or substitutions with disease severity, these findings underscore the importance of reporting genetic surveillance data along with epidemiological data to allow for analysis of factors that may increase risk of influenza and impact disease severity [32]. Although we did not observe association between viral load and disease severity, larger studies have reported this association among hospitalized patients with pneumonia infection [33], which underscores the need for multi-site studies with improved statistical power to estimate associations. We reported a reduction in disease severity with increase in age in children aged <5 years, which corroborates the evidence that children aged <6 months may experience more severe influenza related complications [34].

The study had some limitations. First, the analysis in this report only involved the HA, NA, M2, and NS1 gene segments of A(H1N1)pdm09 virus. Although these regions are important in understanding antigenic drift and antiviral drug resistance in influenza viruses, important changes in other gene segments, for example, mutations associated with increased pathogenicity may not have been captured. Secondly, the prioritized samples were selected based on anticipated probability of successful sequencing inferred from the sample’s viral load as indicated by the diagnosis Ct value. Such a strategy ultimately avoided NGS of some samples that may have been critical in inferring additional genetic characteristics of circulating influenza viruses. Lastly, the analysis in this report did not include phenotypic analyses to assess the effect of observed substitutions on virulence, pathogenicity, and transmissibility of influenza viruses.

In conclusion, our study highlights the utility of genomic surveillance to monitor the evolutionary changes of influenza viruses within a country and the utility of combined genetic and epidemiological data to improve understanding of influenza season severity and guide intervention. Routine influenza surveillance with broad geographic representation and whole genome sequencing capacity to inform on the severity of circulating strains could improve selection of influenza strains for inclusion in vaccines.

## Data Availability

We deposited all the generated sequence data in the Global Initiative on Sharing All Influenza Data (GISAID) EpiFluTM database (https://platform.gisaid.org/epi3/cfrontend).

## Supporting information Conflict of interest

None

## References

1. Garten, R.J., et al., Antigenic and genetic characteristics of swine-origin 2009 A(H1N1) influenza viruses circulating in humans. Science, 2009. 325(5937): p. 197–201.

2. Dawood, F.S., et al., Emergence of a Novel Swine-Origin Influenza A (H1N1) Virus in Humans. New England Journal of Medicine, 2009. 360(25): p. 2605–2615.

3. Smith, G.J., et al., Origins and evolutionary genomics of the 2009 swine-origin H1N1 influenza A epidemic. Nature, 2009. 459(7250): p. 1122–5.

4. Baillie, G.J., et al., Evolutionary dynamics of local pandemic H1N1/2009 influenza virus lineages revealed by whole-genome analysis. J Virol, 2012. 86(1): p. 11–8.

5. Zehender, G., et al., Reconstruction of the evolutionary dynamics of the A(H1N1)pdm09 influenza virus in Italy during the pandemic and post-pandemic phases. PLoS One, 2012. 7(11): p. e47517.

6. Venter, M., et al., Evolutionary dynamics of 2009 pandemic influenza A virus subtype H1N1 in South Africa during 2009-2010. J Infect Dis, 2012. 206 Suppl 1: p. S166–72.

7. Katz, M.A., et al., Results From the First Six Years of National Sentinel Surveillance for Influenza in Kenya, July 2007–June 2013. PLoS ONE, 2014. 9(6): p. e98615.

8. Emukule, G.O., et al., The burden of influenza and RSV among inpatients and outpatients in rural western Kenya, 2009-2012. PLoS One, 2014. 9(8): p. e105543.

9. Emukule, G.O., et al., Influenza activity in Kenya, 2007-2013: timing, association with climatic factors, and implications for vaccination campaigns. Influenza and Other Respiratory Viruses, 2016. 10(5): p. 375–385.

10. Lemey, P., M. Suchard, and A. Rambaut, Reconstructing the initial global spread of a human influenza pandemic: a Bayesian spatial-temporal model for the global spread of H1N1pdm. PLoS Curr Biol, 2009. 1(RRN1031).

11. Rambaut, A. and E. Holmes, The early molecular epidemiology of the swine-origin A/H1N1 human influenza pandemic. PLoS Currents, 2009. 1: p. RRN1003.

12. Nokes, D.J., et al., Incidence and severity of respiratory syncytial virus pneumonia in rural Kenyan children identified through hospital surveillance. Clin Infect Dis, 2009. 49(9): p. 1341–9.

13. Owuor, D.C., et al., Characterizing the Countrywide Epidemic Spread of Influenza A(H1N1)pdm09 Virus in Kenya between 2009 and 2018. Viruses, 2021. 13(10): p. 1956.

14. Katz, M.A., et al., Epidemiology, Seasonality, and Burden of Influenza and Influenza-Like Illness in Urban and Rural Kenya, 2007-2013;2010. The Journal of Infectious Diseases, 2012. 206: p. S53–S60.

15. Feikin, D.R., et al., The burden of common infectious disease syndromes at the clinic and household level from population-based surveillance in rural and urban Kenya. PLoS One, 2011. 6(1): p. e16085.

16. Emukule, G.O., et al., The Epidemiology and Burden of Influenza B/Victoria and B/Yamagata Lineages in Kenya, 2012-2016. Open forum infectious diseases, 2019. 6(10): p. ofz421–ofz421.

17. Zhou, B. and D.E. Wentworth, Influenza A virus molecular virology techniques. Methods Mol Biol, 2012. 865: p. 175–92.

18. A.J., C., et al., The antigenic structure of the influenza virus A/PR/8/34 hemagglutinin (H1 subtype). Cell, 1982. 31(2 Pt 1): p. 417–427.

19. Nguyen, H.T., A.M. Fry, and L.V. Gubareva, Neuraminidase inhibitor resistance in influenza viruses and laboratory testing methods. Antivir Ther, 2012. 17(1 Pt B): p. 159–73.

20. Clark, A.M., et al., Functional Evolution of Influenza Virus NS1 Protein in Currently Circulating Human 2009 Pandemic H1N1 Viruses. J Virol, 2017. 91(17).

21. Gachara, G., et al., Whole genome characterization of human influenza A(H1N1)pdm09 viruses isolated from Kenya during the 2009 pandemic. Infect Genet Evol, 2016. 40: p. 98–103.

22. Opanda, S., et al., Assessing antigenic drift and phylogeny of influenza A (H1N1) pdm09 virus in Kenya using HA1 sub-unit of the hemagglutinin gene. PLoS One, 2020. 15(2): p. e0228029.

23. Monamele, C.G., et al., Molecular characterization of influenza A(H1N1)pdm09 in Cameroon during the 2014-2016 influenza seasons. PLoS One, 2019. 14(1): p. e0210119.

24. Dia, N., et al., A subregional analysis of epidemiologic and genetic characteristics of influenza A(H1N1)pdm09 in Africa: Senegal, Cape Verde, Mauritania, and Guinea, 2009-2010. Am J Trop Med Hyg, 2013. 88(5): p. 946–53.

25. Byarugaba, D.K., et al., Whole-genome analysis of influenza A(H1N1)pdm09 viruses isolated in Uganda from 2009 to 2011. Influenza Other Respir Viruses, 2016. 10(6): p. 486–492.

26. Mukherjee, A., et al., Genetic Characterization of Circulating 2015 A(H1N1)pdm09 Influenza Viruses from Eastern India. PLoS One, 2016. 11(12): p. e0168464.

27. Jones, S., et al., Evolutionary, genetic, structural characterization and its functional implications for the influenza A (H1N1) infection outbreak in India from 2009 to 2017. Sci Rep, 2019. 9(1): p. 14690.

28. Dawa, J., et al., Developing a seasonal influenza vaccine recommendation in Kenya: Process and challenges faced by the National Immunization Technical Advisory Group (NITAG). Vaccine, 2019. 37(3): p. 464–472.

29. Owuor, D.C., et al., Genetic characterization of influenza A(H3N2) viruses circulating in coastal Kenya, 2009-2017. Influenza and Other Respiratory Viruses, 2020. 14(3): p. 320–330.

30. Nyasimi, F.M., et al., Epidemiological and evolutionary dynamics of influenza B virus in coastal Kenya as revealed by genomic analysis of strains sampled over a single season. Virus Evolution, 2020. 6(2).

31. Goldstein, E.J., et al., Integrating patient and whole-genome sequencing data to provide insights into the epidemiology of seasonal influenza A (H3N2) viruses. Microbial Genomics, 2017. 4.

32. Broberg, E., et al., Improving influenza virological surveillance in Europe: strain-based reporting of antigenic and genetic characterisation data, 11 European countries, influenza season 2013/14. Euro Surveill, 2016. 21(41).

33. Feikin, D.R., et al., Is Higher Viral Load in the Upper Respiratory Tract Associated With Severe Pneumonia? Findings From the PERCH Study. Clin Infect Dis, 2017. 64(Suppl_3): p. S337–S346.

34. Lafond, K.E., et al., Global Role and Burden of Influenza in Pediatric Respiratory Hospitalizations, 1982-2012: A Systematic Analysis. PLoS medicine, 2016. 13(3): p. e1001977–e1001977.

